# Population-based Cross-sectional Analysis of the Epidemiology of the Surgical Correction of Hyperhidrosis in 1216 Patients over 11 years

**DOI:** 10.1101/2021.09.14.21263594

**Authors:** Marcelo Fiorelli Alexandrino da Silva, Andressa Cristina Sposato Louzada, Marcelo Passos Teivelis, Nickolas Stabellini, Dafne Braga Diamante Leiderman, José Ribas Milanez de Campos, Edson Amaro, Nelson Wolosker

## Abstract

**Background:** Endoscopic thoracic sympathectomy is the definitive surgical treatment for hyperhidrosis, and a nationwide study suggested that it has been performed in a higher rate than which could have been expected due to climate characteristics, comparing to the national statistics.

**Objectives:** To study the epidemiology of sympathectomy to treat hyperhidrosis in São Paulo, the largest city in the Southeast.

**Design and setting:** population-based cross-sectional study.

**Methods:** data on sympathectomies to treat hyperhidrosis between 2008 and 2018 were assessed from the database of the Municipal Health Secretary of São Paulo, Brazil.

**Results:** 65.29% of the patients were female, 66.2% aged between 20 and 39 years and 37.59% had addresses registered outside São Paulo. 1216 procedures were performed in the city of São Paulo from 2008 to 2018, 78.45% of which in only two public hospitals. The number of procedures significantly declined over the years (p = 0.001). 71.63% of the procedures were associated with 2 to 3 days of hospital stay, only 78 intensive care unit days were billed and we did not observe any intra-hospital death.

**Conclusions:** sympathectomies for the treatment of HH were widely performed in the city of São Paulo (1216 procedures), mainly in young (20 -39 years) and female patients, with more than one third of the patients having addresses registered outside the city. This is a very safe surgery, with low need for intensive care units and no mortality in our series. There was a decreasing trend in the number of surgeries over the years.

## Introduction

Sweat exceeding the needs of thermoregulation in certain body areas due to hyperfunctioning sweat glands characterizes hyperhidrosis (HH), which seems to be related to higher levels of cholinergic acetylcholine and nicotinic alpha-7 receptors in their sympathetic ganglia(1). This disease, which affects up to 2.8-4.8% of the population(2,3), has a significant negative impact on patients’ quality of life, impairing personal and professional relationships(4). HH commonly begins in childhood and can persist throughout adulthood if not properly treated(2,5).

Currently, the definitive surgical treatment for HH consists of endoscopic thoracic sympathectomy (ETS), which is safe and clinically effective, resulting in significant improvement in quality of life(4,6,7). Although this treatment is covered by the Brazilian Public Health System (SUS), a recent nationwide analysis reported great variability in regional rates of ETS to treat HH that could not be explained by climatic differences(8). For instance, in this study, the Southeast region accounted for almost half of the procedures, corresponding to the second highest standardized procedure rate per inhabitants per year among the 5 Brazilian regions(8), even though it is the second coldest region and also not the wettest(9). On the other hand, the Southeast has the best developmental indexes and greater physician density(9). Hence, we hypothesized that cultural and socioeconomic factors have contributed to the numbers observed in the Southeast and perhaps the epidemiology and outcomes of ETS to treat HH are different from those in Brazil.

Therefore, we designed the present study to assess the epidemiology and outcomes of ETS to treat HH in the city of São Paulo, which is the largest city of the Southeast, with an estimated population of 12 million inhabitants(9), with more than 5 million exclusively dependent on SUS(10). Added to that, the database of the Municipal Health Secretary of São Paulo provides the most detailed health data(11), yielding more information than the national database, such as hospital volumes, hospital and intensive care unit (ICU) length of stay, as well as the proportion of patients from other cities who underwent ETS to treat HH in São Paulo, which might shed light on a possible health care tourism.

## Methods

This study was conducted with analysis of data available on the TabNet platform of the DATASUS system(11), which provides open data on procedures performed in accredited public hospitals. Such accreditation is a prerequisite for government reimbursement for the surgeries performed.

Data regarding sympatectomies for the treatment of HH (as coded according to the International Classification of Diseases, 10^th^ edition: R61) were selected from 2008 to 2018 in TabNet of the Municipal Health Department of São Paulo. Among the selections, sex, municipality of residence, age group, number of surgeries performed (total and per establishment), mortality during hospitalization, length of stay in the establishment, intensive care unit (ICU) stay and paid amounts were analyzed.

Age was stratified as follows: under 14 years, between 15 and 19 years, between 20 and 39 years and over 39 years. The amounts paid in reais (Brazilian official currency) were converted into US dollars on December 31, 2012 (which is the intermediate date between the first and last data analyzed, 1USD=R$2.04). Hospitals were numbered, in descending order, by the total number of procedures performed.

The information was obtained from publicly accessible websites by means of computer programs for accessing content from web scraping codes. These codes were programmed in Python language (v. 2.7.13, Beaverton - Oregon - USA) on the Windows 10 Single Language operating system. The steps of collecting and selecting fields on the platform and later adjusting the tables were performed using the selenium-webdriver packages (v. 3.1.8, Selenium HQ, various contributors around the world) and pandas (v. 2.7.13, Lambda Foundry, Inc. and PyData Development Team, New York, USA). We used the browser Mozilla Firefox (v. 59.0.2, Mountain - California - USA) and webdriver geckodriver (v 0.18.0, Mozilla Corporation, Bournemouth, England).

After collection and treatment, data were organized and grouped in a spreadsheet in the Microsoft Office Excel 2016® program (v. 16.0.4456.1003, Redmond - Washington - USA).

For statistical analysis, linear regression analysis was performed to evaluate the trends in procedures over the years, using the SPSS® (IBM Corp. Released 2013. IBM SPSS v 22.0, Armonk, NY – USA). For all tests, the level of statistical significance was α = 0.05.

This study was approved by the Ethics Committee of the Hospital Israelita Albert Einstein. Data are anonymous at DATASUS, therefore a waiver of informed consent was requested and granted by our institutional review board (process number 3067-17).

## Results

Most of the patients were female (65.29%), aged between 20 and 39 years (66.2%) and with a registered residence address in the city of São Paulo (62.41%). The age stratification of the operated patients is presented in table 1.

**Table 1:**
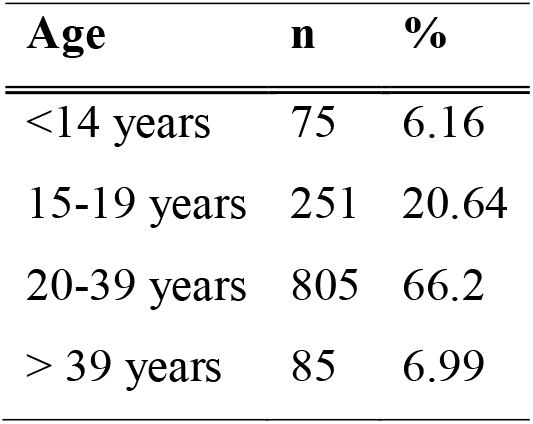
Age stratification divided into 4 subgroups (<14 years, 15-19 years, 20-39 years and> 39 years).

| Age | n | % |
| --- | --- | --- |
| <14 years | 75 | 6.16 |
| 15-19 years | 251 | 20.64 |
| 20-39 years | 805 | 66.2 |
| > 39 years | 85 | 6.99 |

In total, 1216 procedures were performed in the city of São Paulo from 2008 to 2018. Most surgeries (78.45%) were performed in only two public hospitals. The distribution of procedures over the years is shown in Figure 1.

**Figure 1:**
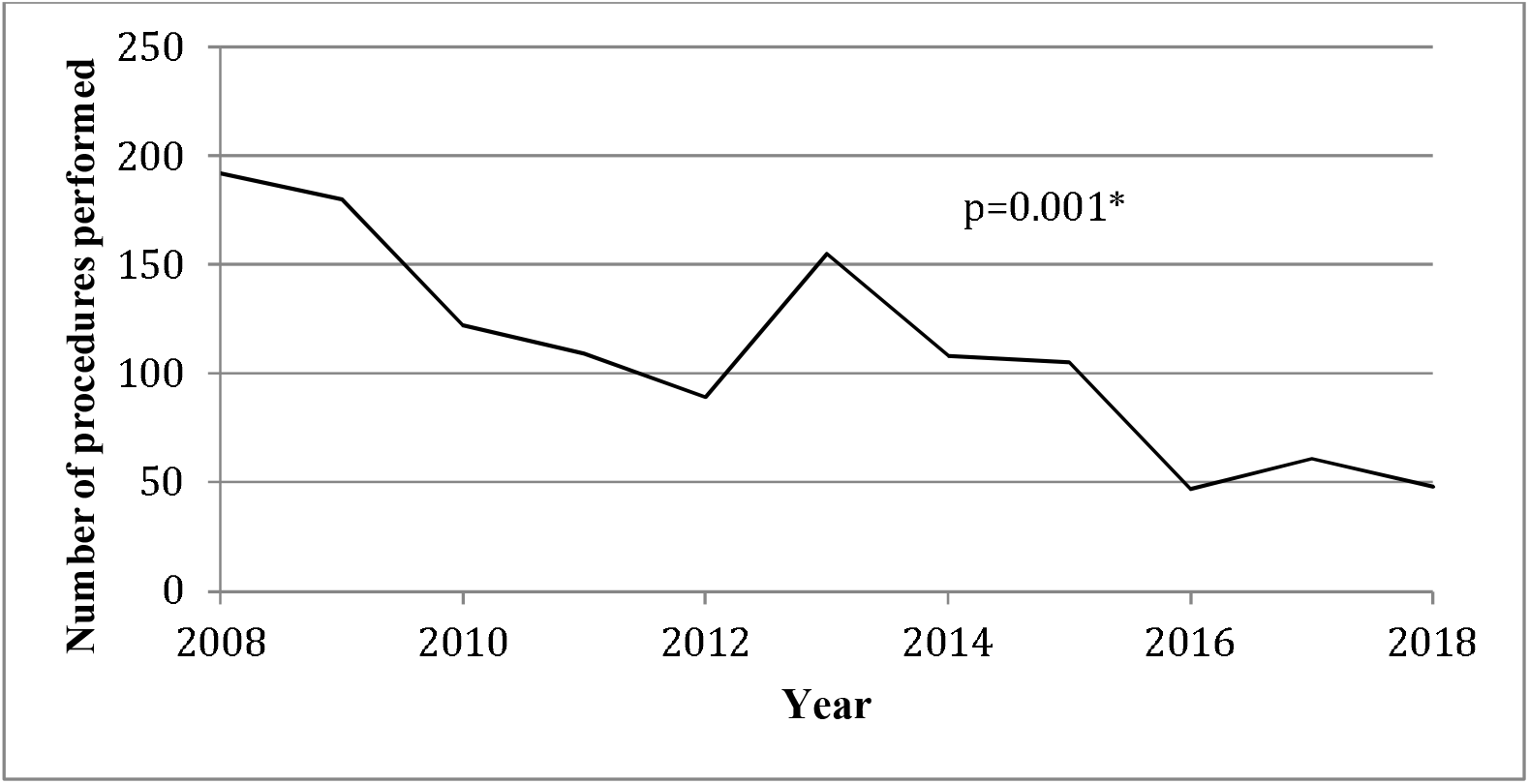
Distribution of sympathectomies for the treatment of hyperhidrosis between 2008 and 2018 in public hospitals in São Paulo. * Linear regression analysis to assess the trend of reducing the number of procedures over the years.

We observed a progressively decreasing trend in the number of procedures over the years (p = 0.001). The first year studied, 2008, presented the largest number of cases (192 surgeries), whereas in recent years, there have been fewer cases, especially in 2016 (47 surgeries).

In all evaluated years, we did not observe any case of intra-hospital death. In other words, no patient died in the same hospital admission after sympathectomy for hyperhidrosis in the city of São Paulo during the studied period.

The number of procedures classified by length of stay is shown in Figure 2.

We observed a predominance of procedures whose length of hospital stay was 2 to 3 days (71.63%). Only 89 patients (7.31%) remained in the hospital for 4 days or more.

**Figure 2:**
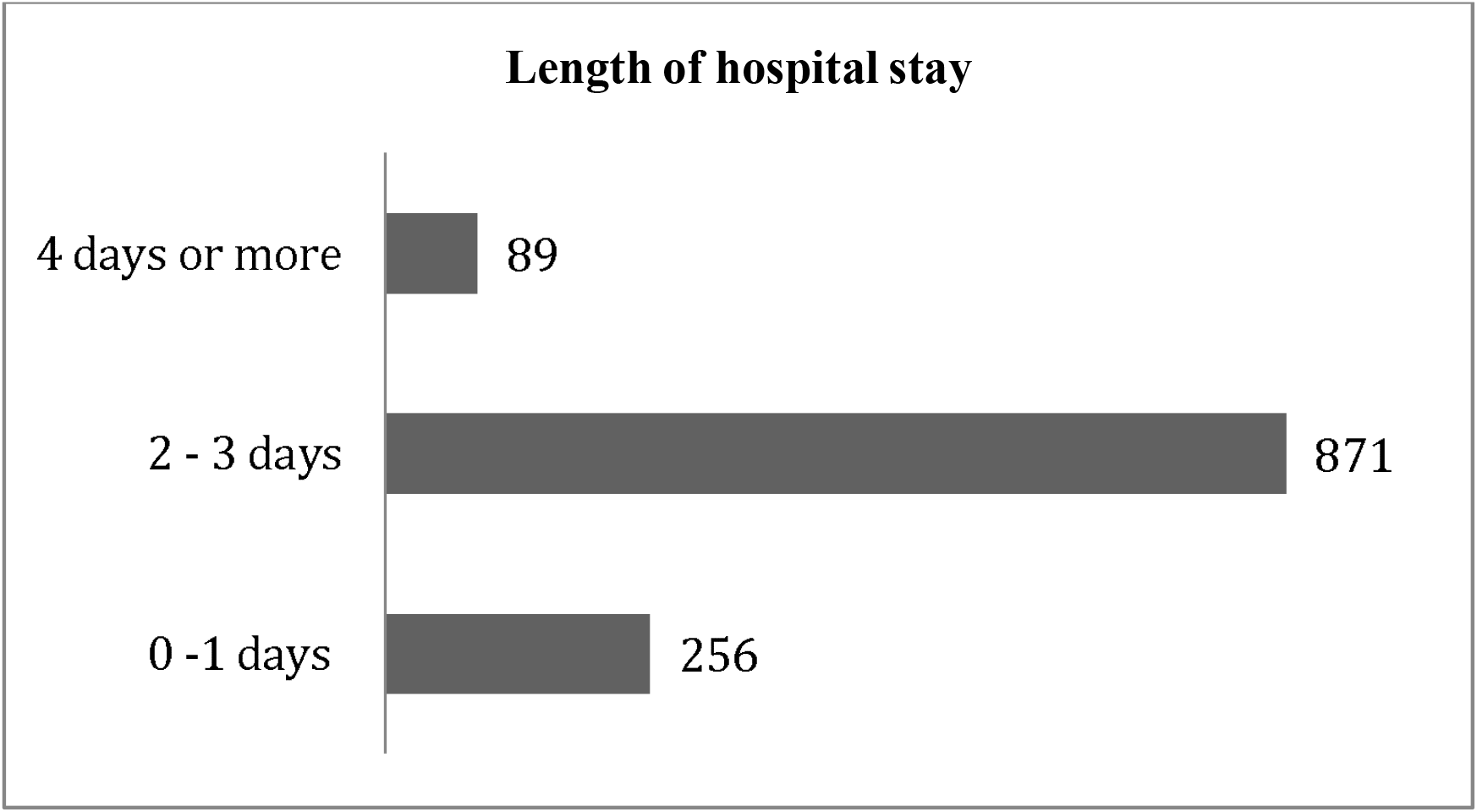
Length of hospital stay in days.

Regarding Intensive Care Unit (ICU) stay, SUS reimbursed a total of 78 days of ICU stay for all cohort: 2 days of ICU were paid for in 2008, 24 days of ICU 2010, 2 days of ICU in 2015 and 50 days of ICU in 2017. In all other years, no ICU stay was recorded for any patient.

The number of procedures, dollar amounts reimbursed by SUS and average amounts by procedure according to hospital establishment are presented in table 2.

**Table 2:** number of procedures (%), values reimbursed by the public health system and average amount per procedure.

| Hospital | Number of procedures (%) | Total reimbursed | Average amount per procedure |
| --- | --- | --- | --- |
| 1 | 753 (61.92) | 314,153.19 | 417.20 |
| 2 | 201 (16.53) | 94,278.25 | 469.04 |
| 3 | 69 (5.67) | 32,493.74 | 470.92 |
| 4 | 66 (5.43) | 33,052.79 | 500.80 |
| 5 | 56 (4.61) | 26,898.84 | 480.34 |
| 6 | 16 (1.32) | 7,610.58 | 475.66 |
| 7 | 11 (0.90) | 5,411.38 | 491.95 |
| 8 | 11 (0.90) | 4,607.50 | 418.86 |
| 9 | 9 (0.74) | 5,467.13 | 607.46 |
| 10 | 9 (0.74) | 4,209.31 | 467.70 |
| 11 | 8 (0.66) | 3,293.05 | 411.63 |
| 12 | 5 (0.41) | 2,336.79 | 467.36 |
| 13 | 1 (0.08) | 658.72 | 658.72 |
| 14 | 1 (0.08) | 445.67 | 445.67 |
| **Total** | **1216 (100)** | **534,916.96** | **439.90** |

A total of $534,916.96 was reimbursed by SUS for all the surgeries, corresponding to an average amount of $439.90 per surgery. The reimbursement per procedure ranged from $417.20 to a maximum of $607.46 throughout the years.

## Discussion

### Patients’ demographics

In line with previous reports, we observed a predominance of female patients(8,12). Even though the prevalence of HH is similar between sexes(5), there is a greater demand for treatment in females, likely due to a generally greater aesthetic concern in this group(13–15).

Regarding age, ETS to treat HH has proven to be beneficial for a wide range of age groups(16,17). However, the demand for treatment of hyperhidrosis is estimated to be greater in young and economically active age groups(5), which is in agreement with our findings, as most individuals in our study were aged between 20 and 39 years (66.2%).

More than one third of the patients had registered addresses outside the city of São Paulo (37.59%). Health care inequalities in Brazil and the concentration of resources in the Southeast, and in some Southeastern centers as the city of São Paulo, are well documented(9). A similar high proportion of out-of-town patients who practice health tourism in order to receive complex treatments in São Paulo has been observed in other reports(18), and makes it inaccurate to calculate a standardized procedure rate per inhabitants per year, as we cannot assess the population quotient.

### Procedure rates and trends

Since the beginning of the study we observed a progressively downward trend in the number of ETS for the treatment of HH, which contrasts with national statistics of an initial upward trend between 2008 and 2012, and a downward trend afterwards(8). This difference may be due to the fact that the municipal center with the highest volume of ETS was also the one in which a “oxybutynin first” protocol for the treatment of HH started in 2007, reserving sympathectomy for refractory cases or for patients with intolerance to the oral treatment, in an attempt to reduce the incidence compensatory hyperhidrosis, a common complication after sympathectomy(19–22). Good results of the “oxybutynin first” strategy have been reported as of 2011(23–28), which might have contributed to the national decrease of ETS performed for the treatment of HH from 2012 onwards.

### Hospital and ICU length of stay and in-hospital mortality

We observed a short hospital stay, in line with other studies(29,30). Most of the patients were discharged on the second or third day of hospitalization, which likely corresponds to the first and second post-operative day, respectively, as they are usually admitted the day before the surgery.

Regarding ICU length of stay, due to the anonymous data, we only have access to the total ICU days paid by the government, we do not know how many patients were admitted in the ICU, for how many days or what were the indications for ICU, which are limitations of our study. In total, we observed 78 days unevenly distributed over the years of 2008, 2010, 2015 and 2017 only. This number is small, but not negligible, especially when considering most of the patients were young adults. Possibly some of these days in the ICU correspond to patients with severe early post-operative complications, such as pneumothorax or hemothorax, which have been reported by other authors(29). Nevertheless, no in-hospital death after ETS to treat HH was reported in the public hospitals of São Paulo between 2008 and 2018, as reported in other large studies(29,31), which emphasizes the low mortality associated with this treatment.

Within the city of São Paulo 78.45% of the procedures were performed in only two public hospitals. As hyperhidrosis is a bothersome, but not lethal disease, and considering the underfunding of the public health system(32), there are few referral centers dedicated to this treatment, which tend to improve peri-operative outcomes, and may likely explain the absence of fatalities.

### Costs

A total of $534,916.96 was reimbursed by the government for ETS to treat HH, with an average amount of $439.90 per procedure, a little less than reported by other authors(33). One of the limitations of our cost analysis is that the reimbursement is based on a compensation table for procedures, which often do not reflect the actual amount spent on procedures by the hospitals. We may conjecture that the decrease in the number of surgeries may lead to higher public expenses for clinical treatments. As HH is a frequent disease in young people, who may use medications for the long term, this should be considered in a publicly funded health care, although a cost-effective study is necessary to evaluate it properly.

### Limitations

Besides the limitations already mentioned, that are, anonymous data precluding follow up and an adjusted analysis, reimbursement based on a fixed compensation table, another limitation of our study is that the database only provides data related to selected procedures performed in accredited public hospitals, being susceptible to some loss of data.

On the other hand, this is a comprehensive and detailed epidemiological analysis of sympathectomy for the treatment of HH, analyzing objective and compulsorily recorded data in the public data. Our findings ensure the demographics of patients who seek surgical treatment for hyperhidrosis, present the high volume of out-of-town patients who seek treatment in São Paulo, and highlight the safety of this treatment.

### Conclusions

In the last 11 years, sympathectomies for the treatment of HH were widely performed in the city of São Paulo (1216 procedures), mainly in young (20 -39 years) and female patients, with more than one third of the patients having addresses registered outside the city. This is a very safe surgery, with low need for intensive care units and no mortality in our series. There was a decreasing trend in the number of surgeries over the years.

## Data Availability

Data is publicly available at datasus

http://tabnet.saude.sp.gov.br/deftohtm.exe?tabnet/aih_rd2008.def

## Notes

### Competing Interest Statement

The authors have declared no competing interest.

### Funding Statement

Nickolas Stabellini (author) received an undergraduate research scholarship from Institutional Scientific Initiation Scholarship Program (PIBIC). Process number 800996 2018 6, granted by the National Council for Scientific and Technological Development (CNPq), Brazil.

### Author Declarations

This study was approved by the Ethics Committee of the Hospital Israelita Albert Einstein. Data are anonymous at DATASUS, therefore a waiver of informed consent was requested and granted by our institutional review board (process number 3067 17).

